# Strengthening Emergency Care Capacity to Increase Inpatient Admissions: A Single-Center Quality Improvement Project in Tokyo

**DOI:** 10.1101/2025.10.26.25338805

**Authors:** Tatsuo Yanagawa, Miki Tamura, Minako Inui, Kazumi Fukumoto, Matsuko Sato, Koichiro Azuma, Naoto Kurihara

**Affiliations:** Department of Medicine, Nerima General Hospital, 1-24-1 Asahigaoka, Nerima Ward, Tokyo 176-8530, Japan; Department of Nursing, Nerima General Hospital, 1-24-1 Asahigaoka, Nerima Ward, Tokyo 176-8530, Japan; Department of Gastrointestinal Surgery, Nerima General Hospital, 1-24-1 Asahigaoka, Nerima Ward, Tokyo 176-8530, Japan; Institute of Healthcare Quality Improvement, Public Interest Incorporated Foundation Tokyo Healthcare Foundation, Tokyo, Japan

**Keywords:** Continuous Quality Improvement, Before–After Study, Emergency Department, Health Services Research, Performance Measures

## Abstract

**Background:** Stabilizing inpatient volume is essential for acute care hospitals to sustain operations and meet community needs. Although Japan overall has a high national bed supply, Nerima Ward in Tokyo has one of the lowest bed-to-population ratios, resulting in limited access to acute care. At our 224-bed hospital, ambulance acceptance and inpatient admissions had remained stagnant despite consistent regional demand, suggesting that limited emergency intake capacity might be constraining overall inpatient volume. Therefore, we implemented a series of coordinated operational improvements aimed at strengthening emergency care capacity and investigated whether increasing ambulance acceptance could effectively raise inpatient admissions and improve access to acute care.

**Methods:** We conducted a single-center, observational before-after study using monthly aggregated data from January 2024 to July 2025. Outcomes included the number of inpatients, ambulance-based admissions, ambulance arrivals, discharges, bed occupancy, length of stay, and surgeries. Changes before and after the operational change were compared using Welch t-test. Multivariable regression identified independent predictors of inpatient volume.

**Results:** After the operational change, the mean monthly number of inpatients increased (+49.3, *p*=0.003), corresponding to approximately a 10% relative increase. Ambulance-based admissions (+29.0, *p*<0.001), ambulance arrivals (+60.1, *p*=0.001), and discharges (+39.6, *p*=0.018) also rose significantly, while bed occupancy was maintained despite shorter stays. Regression analysis identified ambulance-based admissions (β=+1.24, *p*<0.001) and the number of surgeries (β=+1.09, *p*=0.002) as independent predictors of inpatient volume (adjusted R squared=0.725).

**Conclusion:** Enhancing emergency care capacity-mainly through increased ambulance acceptance combined with other coordinated measures-substantially increased inpatient volume and improved access to acute care. These findings underscore the importance of integrated, multi-faceted operational improvements over single-focus interventions for hospital sustainability.

**Key Messages:** *What is already known on this topic:* In Japan, planned and referral-based admissions dominate, while the contribution of emergency transports to inpatient volume remains underexplored.

*What this study adds:* Strengthening overall emergency care capacity, including triage coordination and surgical throughput, led to a sustained increase in inpatient volume.

*How this study might affect research, practice or policy:* Integrated emergency and surgical capacity building may enhance timely patient access and support the sustainability of acute care hospitals in bed-limited regions.

## Introduction

Stabilizing inpatient volume is a critical challenge for acute care hospitals, both for ensuring sustainable operations and for meeting community healthcare needs. Internationally, the pathways leading to hospitalization vary widely across countries. In the United States, emergency departments serve as the primary gateway to inpatient care [1], and in the United Kingdom, the Accident & Emergency (A&E) department similarly functions as the main route for emergency admissions [2].

In contrast, in Japan, visits to the emergency department do not necessarily lead to hospitalization. According to the Patient Survey 2020 by the Ministry of Health, Labour and Welfare, only about 20–25% of hospitalizations occur through the emergency department [3], indicating that emergency-based admissions represent a smaller proportion of total hospitalizations in Japan compared with Western countries.

Therefore, these findings suggest that in Japan, the emergency department plays a relatively limited role as a pathway to hospitalization.Japan’s hospital bed supply is exceptional, averaging 8.1 acute care beds per 1,000 population [4]—more than twice the OECD average of 3.5. This abundance reflects structural characteristics such as slower transitions to outpatient and home-based care and challenges in coordinating post-discharge services. However, this national average conceals striking regional variation [5].In Tokyo, certain urban wards such as Nerima represent a contrasting context. Despite being densely populated, these areas maintain a relatively low supply of acute care beds compared with the national average, reflecting the uneven distribution of healthcare resources across Japan. The population is gradually increasing, with 21.6% aged 65 years or older, while also experiencing an influx of residents in their 30s. These demographic dynamics generate rising healthcare demand, particularly for emergency and acute services, while the younger cohorts modestly support workforce availability.

Our hospital, a 224-bed acute care facility located in eastern Nerima, is the only mid-sized hospital in the area. Larger tertiary centers (>400 beds) are concentrated in the western part of the ward, creating an uneven distribution of healthcare resources. Consequently, a substantial number of emergency and inpatient cases have been diverted outside the ward, despite persistent community demand for acute care.

However, our ambulance acceptance capacity had reached its operational limit due to staffing constraints, limited inpatient coordination, and competition for beds. This mismatch between community demand and hospital capacity led to stagnation in both ambulance acceptance and inpatient admissions, raising concerns for regional access to emergency care and hospital sustainability.

In Japan, the Diagnosis Procedure Combination (DPC) system—a case-mix–based reimbursement framework similar to Diagnosis-Related Groups (DRGs)—was introduced to improve transparency, efficiency, and quality in acute inpatient care [6]. Under this system, hospitals are incentivized to treat acute, time-sensitive cases efficiently, particularly those requiring emergency admission or surgical intervention. Understanding the operational factors that drive inpatient volume within this context is therefore essential for aligning clinical capacity with both community needs and financial viability.

To address these challenges, we initiated a structured quality improvement (QI) project in October 2024 aimed at strengthening emergency care capacity and improving patient flow. The objectives of this study were: (1) to evaluate whether the operational intervention increased the number of inpatients, and (2) to identify the primary determinants of inpatient volume in this context.

## Methods

### Study setting

This study was conducted at a 224-bed acute care hospital in eastern Nerima Ward, Tokyo. The hospital provides 19 specialties and is the sole mid-sized acute care facility in this part of the ward, serving a mixed population of aging residents and younger in-migrants. At baseline, the hospital received approximately 330 ambulance cases per month—around 30 fewer than the operational target—reflecting limitations in emergency acceptance capacity. Frequent ambulance diversions occurred, and some emergency requests had to be declined due to staffing and bed availability constraints. Consequently, limited ambulance acceptance and stagnant inpatient admissions represented pressing operational issues.

### Operational change

In October 2024, a series of quality improvement measures was introduced to strengthen emergency care capacity. Key initiatives included: reorganization of emergency nursing roles, restructuring of physician on-call schedules to ensure 24-hour coverage, reinforcement of bed management through multidisciplinary meetings [7], direct handling of ambulance service calls by the charge nurse in the emergency department, expansion of night-shift laboratory and radiology support (urgent blood tests added in January 2025) [8], recruitment of a neurosurgeon (April 2024), and additional measures such as removal of infection-control tents, task shifting in radiology, enhanced nursing education, and revised weekend device management. The charge nurse also assumed responsibility for managing the emergency communication terminal throughout the day, consistent with prior literature describing the charge nurse’s central role in coordinating information flow and communication in the emergency department [9,10]. In Japan, this terminal system allows hospitals to indicate when they cannot accept new patients, enabling ambulance crews to identify available hospitals in real time. [11]

### Study design

We conducted a single-center observational before–after study using monthly aggregated data from January 2024 to July 2025 (19 months). The study exploited a natural experiment, as the hospital independently implemented the operational change to strengthen emergency acceptance. This study was reported in accordance with the SQUIRE 2.0 guidelines (Standards for Quality Improvement Reporting Excellence).

### Data collection and measures

Data were obtained from the hospital’s administrative records. Eleven operational indicators were collected: number of inpatients, total inpatient days, ambulance-based admissions, number of ambulance arrivals, number of discharges, bed occupancy rate, average length of stay, number of surgeries, total outpatient visits, number of new outpatients, and number of referral letters.

In the present study, the number of inpatients was defined as the dependent variable, while the remaining ten indicators were considered as candidate explanatory variables for multivariable analyses. Previous studies have frequently used related variables—such as ambulance transport, length of stay, or bed occupancy rate—to analyze hospital admissions or admission probabilities rather than absolute inpatient counts [12–15]. In contrast, this study focused on inpatient volume itself, aiming to extend prior approaches. These indicators were used for two objectives: (1) to examine whether the operational change was associated with variations in outcomes—particularly inpatient numbers, ambulance arrivals, and ambulance-based admissions; and (2) to identify determinants of inpatient volume through statistical analyses.

### Statistical analysis

Monthly data were summarized as means with standard deviations. Values before and after the operational change (January–September 2024 vs October 2024–July 2025) were compared using Welch’s t-test (Table 1). For regression analysis, explanatory variables were screened using variance inflation factors (VIF) and Pearson’s correlation coefficients (Table 2). Variables with VIF ≥10 or without significant correlation with the number of inpatients were excluded. The final regression model included the remaining variables. Analyses were conducted using SPSS Statistics version 28.0 (IBM Corp., Armonk, NY, USA), with statistical significance defined as two-sided p<0. 05..

**Table 1.** Comparison of hospital performance indicators before and after the operational change (mean ± SD, Welch’s t-test)

| Indicator | Unit | Before operational change<br>2024.1-2024.9 | After operational change<br>2024.10-2025.7 | values before and after the operational change | p value |
| --- | --- | --- | --- | --- | --- |
| Number of inpatients | patients/mo | 496.8 ± 31.1 | 546.1 ± 30.2 | 49.3 | 0.0028 |
| Total inpatient days | patients/mo | 4957.7 ± 211.7 | 5025.8 ± 252.2 | 68.1 | 0.531 |
| Ambulance-based admissions | patients/mo | 90.2 ± 10.8 | 119.2 ± 10.7 | 29 | <0.001 |
| Number of ambulance arrivals | amb/mo | 334.8 ± 24.7 | 394.9 ± 37.6 | 60.1 | <0.001 |
| Number of discharges | patients/mo | 477.6 ± 34.8 | 517.2 ± 30.6 | 39.6 | 0.0183 |
| Bed occupancy rate | % | 80.0 ± 3.2 | 81.4 ± 3.6 | 1.4 | 0.381 |
| Average length of stay | day | 10.4 ± 0.8 | 9.7 ± 0.7 | -0.7 | 0.073 |
| Number of surgeries | cases/mo | 260.4 ± 18.0 | 275.8 ± 14.1 | 15.4 | 0.057 |
| Total outpatient visits | patients/mo | 9600.3±435.2 | 10011.3±477.1 | 411 | 0.0661 |
| Number of new outpatients | patients/mo | 1352.3±94.1 | 1373.9±82.5 | 21.6 | 0.604 |
| Number of referral letters | letter/mo | 527.2±47.4 | 571.0±56.4 | 43.8 | 0.0837 |

Data are presented as mean ± standard deviation (SD). Each indicator was compared before and after the operational change using Welch's t-test. Units are expressed as patients per month (patients/mo), ambulances per month (amb/mo), surgeries per month (cases/mo), referral letters per month (letters/mo), percentage (%), and days.

**Table 2.** Variance inflation factors (VIF) and Pearson correlation coefficients (r) for independent variables predicting inpatient volume.

| Independent variables | VIF | Pearson correlation coefficients |  |  |
| --- | --- | --- | --- | --- |
|  |  | r | tvalue | P value |
| Total inpatient days | 112.89 | 0.48 | 2.23 | 0.039 |
| Ambulance-based admissions | 6.86 | 0.74 | 4.53 | <0.001 |
| Number of ambulance arrivals | 5.54 | 0.61 | 3.18 | 0.005 |
| Number of discharges | 251.4 | 0.76 | 4.87 | <0.001 |
| Bed occupancy rate | 5.94 | 0.3 | 1.3 | 0.211 |
| Average length of stay | 268.08 | -0.44 | -2.02 | 0.06 |
| Number of surgeries | 7.75 | 0.68 | 3.84 | 0.001 |
| Total outpatient visits | 12.41 | 0.74 | 4.48 | <0.001 |
| Number of new outpatients | 4.68 | 0.48 | 2.25 | 0.038 |
| Number of referral letters | 4.59 | 0.64 | 3.43 | 0.003 |

This table presents the variance inflation factors (VIF) and Pearson's correlation coefficients (r) for each independent variable with respect to the dependent variable (number of inpatients). Independent variables with a $VIF \geq 10$ or a non-significant correlation (r) were excluded. This process demonstrates the rationale for variable selection prior to conducting the multiple regression analysis.

## Results

### Comparison of indicators before and after the operational change

Table 1 shows the monthly means of 11 performance indicators before and after the operational change. Significant increases were observed in the number of inpatients (+49.3 patients/month, p=0.0028), ambulance-based admissions (+29.0 patients/month, p<0.001), ambulance arrivals (+60.1/month, p<0.001), and discharges (+39.6/month, p=0.0183).

Before the operational change, the hospital received approximately 335 ambulance arrivals per month—around 30 below the operational target of 360. After the operational change, this number rose by about 60 to an average of 395 arrivals per month, consistent with the statistically significant increase shown in Table 1 (+60.1/month, p < 0.001). Other indicators, including bed-occupancy rate, average length of stay, total outpatient visits, and number of referral letters, showed no significant change.

### Multiple regression analysis

To identify determinants of number of inpatients, independent variables were evaluated for collinearity using VIF and Pearson’s correlation (Table 2). After excluding variables with VIF ≥10 or non-significant correlations, ambulance-based admissions and number of surgeries were retained in the final model. Multiple regression analysis demonstrated that both ambulance-based admissions (β=+1.24, p<0.001) and number of surgeries (β=+1.09, p=0.002) were independently associated with number of inpatients, with an adjusted R^2^ of 0.725, indicating substantial explanatory power of the model. Model diagnostics indicated no major violations of linear regression assumptions, including normality, independence, and homoscedasticity of residuals.

## Discussion

This quality improvement project demonstrated that enhancing emergency care capacity led to a meaningful increase in inpatient admissions. Importantly, the operational change did not create new demand but rather enabled existing emergency demand to be met more effectively by addressing operational bottlenecks such as triage delays, inpatient flow constraints, and coordination with surgical units. The main contributors to this increase were ambulance-based admissions and the number of surgeries, indicating that improvements in both emergency responsiveness and surgical throughput jointly drove the rise in inpatient volume. This finding is consistent with previous evidence suggesting that hospitals’ ability to convert emergency demand into admissions and maintain timely surgical care are critical determinants of overall utilization [16,17].

Outpatient-related indicators, such as total outpatient visits and referral letters, were not significant predictors, suggesting that, in the short term, inpatient growth stems primarily from emergency and surgical pathways rather than ambulatory channels. The observed ~10% increase in inpatient volume is operationally meaningful for a 224-bed hospital, helping to stabilize resource use and strengthen financial viability. Given that nearly 70% of acute care hospitals in Japan operate at a deficit, even modest improvements in occupancy and throughput can substantially enhance sustainability.

The maintenance of high bed occupancy despite a shorter average stay suggests that capacity was used more efficiently. This improvement likely reflects enhanced coordination between emergency and inpatient units, facilitating smoother patient flow. Consistent with previous reports, structured bed management and multidisciplinary collaboration can mitigate emergency department crowding while maintaining hospital throughput [18]. Few previous studies have directly examined inpatient counts as the main outcome. By focusing on actual patient numbers rather than derived indicators such as admission rates or bed occupancy, this study treats inpatient volume as a concrete measure of hospital performance that reflects the real capacity to meet community demand. In doing so, the analysis provides a simple yet meaningful metric that can be readily applied to evaluate operational performance in similar settings.

## Regional context

Nerima Ward’s inpatient bed supply is less than half of the Tokyo average, representing a striking shortage relative to local population needs. At the same time, aging and moderate in-migration have increased healthcare demand, especially for emergency transport. Strengthening emergency care capacity in such settings is thus essential not only to expand access but also to maintain operational stability for community hospitals.

## Policy and clinical implications

The findings illustrate that aligning emergency capacity with regional bed supply disparities can improve both community access and hospital sustainability. Japan’s DPC (Diagnosis Procedure Combination) system—analogous to diagnosis-related groups (DRGs)—reimburses hospitals based on case mix and efficiency, thereby rewarding effective management of acute emergency and surgical cases. In regions like Nerima Ward, where these patient categories predominate, enhancing ambulance acceptance and surgical capability directly fulfills policy goals and supports hospital finances. This dual alignment between clinical demand and reimbursement structure provides a framework for sustaining acute-care hospitals in resource-limited urban areas. Beyond economic effects, improved emergency access also reinforces public trust and timely care for time-sensitive conditions such as stroke and trauma.

Although the DPC framework is unique to Japan, the broader principle—linking emergency responsiveness to both efficiency and sustainability—has international relevance. Nonetheless, these results should be interpreted cautiously, given the single-center design and urban context.

## Quality improvement perspective

From a quality improvement standpoint, this initiative demonstrates that targeted, system-level interventions—rather than isolated clinical changes—can yield measurable organizational benefits. Comparable efforts, such as those by Katayama et al. [19] in Osaka and Marsilio et al. [20] in Europe, have likewise shown that redesigning emergency pathways can enhance throughput and operational efficiency, while fostering greater staff engagement within improvement initiatives. Our experience suggests that operational gains that are visible to staff not only enhance performance metrics but also strengthen teamwork and morale, which are critical for sustaining a QI culture. Future projects should extend evaluation to include patient outcomes, safety, and satisfaction to achieve a more comprehensive assessment of effectiveness.

## Limitations

This study was limited to a single institution and a 19-month period, which may not fully capture seasonal or long-term variations. Data were aggregated monthly, preventing patient-level analyses. Clinical outcomes such as mortality and safety indicators were not assessed. Future multi-center studies with longer observation periods and patient-level outcome data are warranted to confirm the generalizability of these findings.

## Conclusion

Enhancing emergency care capacity through coordinated operational improvements can effectively increase inpatient volume, improve hospital sustainability, and expand community access to acute care. This approach represents a scalable model for urban hospitals facing resource constraints.

## Data Availability

The data that support the findings of this study are derived from the administrative records of Nerima General Hospital. Due to institutional privacy regulations, these data are not publicly available but can be shared by the corresponding author upon reasonable request and with permission from the hospital.

## Footnotes

### Contributors

MT, MI, KF, MS, KA, and NK discussed and implemented the improvements in emergency operations. MT and MI prepared the procedural manual for the interventions. TY, KA, and NK collected the data. TY drafted the manuscript, with critical input and revisions from KA and NK. All authors critically revised the manuscript for important intellectual content and approved the final version. TY serves as the guarantor. Generative artificial intelligence (ChatGPT, version 5.0; OpenAI) was used to assist in translating the text into academic English.

### Funding

This research received no specific grant from any funding agency in the public, commercial, or not-for-profit sectors.

### Competing interests

None declared.

### Patient and public involvement

Patients and/or the public were not involved in the design, conduct, reporting, or dissemination plans of this research.

### Provenance and peer review

Not commissioned; not peer reviewed (preprint).

### Data availability statement

The datasets used and analysed during the current study are available from the corresponding author on reasonable request.

### Patient consent for publication

Not applicable.

### Ethics approval

This study analysed aggregated monthly hospital data without access to individual patient information. The Institutional Review Board of Nerima General Hospital reviewed the protocol and confirmed that the use of aggregated hospital-level data without individual identifiers did not require formal ethics approval under national guidelines.

## Acknowledgements

The authors thank Emi Mouri, Nerima General Hospital, for conducting the statistical analyses.

## Notes

### Competing Interest Statement

The authors have declared no competing interest.

### Funding Statement

This study did not receive any funding.

### Author Declarations

The Ethics Committee of Nerima General Hospital waived ethical approval for this work. The study used de-identified aggregated hospital administrative data and did not involve individual informed consent.

## References

1. Pines JM, Hilton JA, Weber EJ, Al Shabanah H, Anderson PD, Bernhard M, et al. International perspectives on emergency department crowding. *Acad Emerg Med*. 2011;18(12):1358–70. doi:10.1111/j.1553-2712.2011.01235.x

2. Peters J, Parry J, Van der Pol M, et al. Emergency hospital admissions via accident and emergency departments in England. J R Soc Med. 2014;107(1):22–28. doi:10.1177/01410768135111423.

3. Ministry of Health, Labour and Welfare. Patient Survey 2020 (in Japanese). Tokyo: MHLW; 2022. Available from: https://www.mhlw.go.jp/toukei/saikin/hw/jyuryo/20/dl/gaikyo-all-g.pdf

4. Ministry of Health, Labour and Welfare (Japan). *Survey of Medical Institutions 2022*. Tokyo: MHLW; 2022.

5. Kobayashi J, Imanaka Y, Noguchi H, Hashimoto H. Factors influencing hospital bed occupancy rate in acute care hospitals: a nationwide analysis in Japan. *J Hosp Manag Health Policy*. 2020;4:14. doi:10.21037/jhmhp.2020.03.03

6. Hayashida K, Murakami G, Matsuda S, Fushimi K. History and profile of Diagnosis Procedure Combination (DPC): development of a real data collection system for acute inpatient care in Japan. J Epidemiol. 2021;31(1):1–11. doi:10.2188/jea.JE20200288.

7. Hammer C, DePrez B, White J, Lewis L, Straughen S, Buchheit R. Enhancing Hospital-Wide Patient Flow to Reduce Emergency Department Crowding and Boarding. J Emerg Nurs. 2022 Sep;48(5):603–609. doi: 10.1016/j.jen.2022.06.002.

8. McKenna P, Heslin SM, Viccellio P, Mallon WK, Hernandez C, Morley EJ. Emergency department and hospital crowding: causes, consequences, and cures. Clin Exp Emerg Med. 2019 Sep;6(3):189–195. doi: 10.15441/ceem.18.022.

9. Holmgren J, Andersson H, Carlström E. Charge nurses’ perceived experience in managing daily work and major incidents in emergency departments: A qualitative study. Int Emerg Nurs. 2022;60:101118. doi:10.1016/j.ienj.2021.101118

10. Singhal N, Maheshwari R, Singh T. What do nurses in charge in emergency departments do? A qualitative study. J Emerg Nurs. 2021;47(5):771–779. doi:10.1016/j.jen.2021.06.002

11. Fukaguchi K, Goto T, Yamamoto T, Yamagami H. Experimental Implementation of NSER Mobile App for Efficient Real-Time Sharing of Prehospital Patient Information With Emergency Departments: Interrupted Time-Series Analysis. JMIR Form Res. 2022 Jul 6;6(7):e37301. doi: 10.2196/37301.

12. Peters J, Noack EM, Loerbroks A. Patients who use emergency medical services have greater hospital admission odds compared to those arriving by other means. PLoS One. 2023;18(7):e0288095. doi:10.1371/journal.pone.0288095

13. Friebel R, Fisher R, Deeny SR, Gardner T, Molloy A, Steventon A. The implications of high bed occupancy rates on readmission rates in England: A longitudinal study. Health Policy. 2019;123(7):765–772. doi:10.1016/j.healthpol.2019.05.006

14. Centers for Medicare & Medicaid Services. The effects of hospital rate-setting programs on volumes, length of stay, and hospital utilization. Health Care Financ Rev. 1983;4(2):47–65. Available from: https://www.cms.gov/files/document/hcfr-4-2-47.pdf

15. Hughes JS, Averill RF, Eisenhandler J, Goldfield NI, Muldoon J, Neff JM. The increasing impact of length of stay outliers on inpatient length of stay: implications for hospital capacity planning. BMC Health Serv Res. 2021;21:1179. doi:10.1186/s12913-021-06972-6

16. McManus ML, Long MC, Cooper A, Litvak E. Variability in surgical caseload and access to intensive care services. *N Engl J Med*. 2003;348(21):2149–56. doi:10.1056/NEJMsa021793

17. Harrison JP, Coppola MN, Wakefield M. The role of hospital capacity, efficiency, and occupancy in hospital admissions. *Health Care Manage Rev*. 2005;30(4):331–9. doi:10.1097/00004010-200510000-00008

18. Hoot NR, Aronsky D. Systematic review of emergency department crowding: causes, effects, and solutions. *Ann Emerg Med*. 2008;52(2):126–36. doi:10.1016/j.annemergmed.2008.03.014

19. Katayama Y, Kitamura T, Kiyohara K, Iwami T, Kawamura T, Izawa J, et al. Improvements in patient acceptance by hospitals following the introduction of a smartphone app for the emergency medical service system: a population-based before-and-after observational study in Osaka City, Japan. *JMIR Mhealth Uhealth*. 2017;5(9):e134. doi:10.2196/mhealth.8190

20. Marsilio M, Roldan ET, Salmasi L, Torrisi G. Operations management solutions to improve ED patient flows: evidence from the Italian NHS. *BMC Health Serv Res*. 2022;22:974. doi:10.1186/s12913-022-08339-x

